# Plasma proteomics improves prediction of recurrent cardiovascular events

**DOI:** 10.64898/2026.04.14.26350861

**Authors:** Yang Liu, Carles Foguet, Chief Ben-Eghan, Elodie Persyn, Matthew Richards, Zongtai Wu, Samuel A Lambert, Adam Butterworth, Angela M Wood, Emanuele Di Angelantonio, Michael Inouye, Scott C Ritchie

## Abstract

**Background and Aims:** Despite treatment, patients with established atherosclerotic cardiovascular disease (ASCVD) are at high risk of recurrent events. Existing clinical risk scores for recurrence provide only moderate predictive performance and rely largely on the same conventional risk factors used to predict disease onset. Proteomics is a promising source of new biomarkers but the technologies need focused use cases in order to achieve utility and implementation. We aimed to determine whether plasma proteomics improves prediction of recurrent cardiovascular events beyond established clinical risk models in secondary prevention in a population-scale cohort.

**Methods:** Plasma proteomic profiles from ∼9,300 participants in the UK Biobank with established ASCVD at baseline were analysed using machine learning methods to derive and evaluate proteomic predictors of recurrent cardiovascular events. The top performing model comprised proteins with non-zero weights (full protein score). Predictive performance of the proteomic predictors, an established clinical risk score (SMART2), and their combination was evaluated across six pre-defined testing datasets representing multiple ethnic and geographic groups. A parsimonious set of proteins with existing clinical-grade enzyme-linked immunosorbent assays (ELISAs) available was then derived.

**Results:** The full protein score achieved higher performance for recurrent ASCVD than the SMART2 risk score across all ethnic and geographic subgroups (mean C-index 0.743 vs 0.653). Adding the full protein score to SMART2 improved discrimination, with the largest increase in White Irish participants (ΔC-index, 0.140; 95% CI, 0.074–0.205; P<0.001). However, adding SMART2 to the protein score provided minimal additional value. The parsimonious score preserved most of the discrimination of the full protein model with C-indices of the recurrent ASCVD risk model comprising age, sex and the parsimonious protein score being nearly identical to the full protein model in the largest testing set (0.723 vs 0.728 for White British in England and Wales). The parsimonious protein score showed a marked gradient of risk with the top, middle and bottom quintiles showing 10-year recurrent ASCVD rates of ∼27.4%, ∼9.6% and ∼2.4%, respectively.

**Conclusions:** In patients with established ASCVD, plasma protein measurements substantially improved prediction of recurrent events beyond conventional clinical risk factors, supporting their potential as a complementary tool to guide secondary prevention of cardiovascular disease.

## Introduction

Despite major advances in management and widespread implementation of guideline-directed medical therapies, recurrent cardiovascular events remain a leading cause of morbidity and mortality worldwide, placing a substantial burden on patients and healthcare systems(1–3). Patients with established atherosclerotic cardiovascular disease (ASCVD) represent a particularly high-risk population, for whom clinical guidelines advocate intensive, lifelong secondary prevention (4–7). Nevertheless, epidemiological studies show a substantial residual risk with approximately 18% of patients experiencing a recurrent event within the first year following an initial event, and more than 30% within five years, even in the context of contemporary treatments (8–10). This persistent burden underscores a critical unmet need for improved risk prediction tools to guide individualised treatment intensity as well as follow-up strategies in secondary prevention (11–13). Current clinical risk scores (e.g. SMART2) provide only moderate predictive performance and rely predominantly on conventional risk factors commonly used in primary prevention settings, including age, blood pressure, and smoking(11). These approaches may not adequately capture the biological drivers of disease progression after an initial event. There is therefore an urgent need to identify and validate novel biomarkers that improve prediction of recurrent risk, better reflect active disease biology, and support more precise secondary prevention.

Plasma proteins reflect dynamic biological processes and can inform cardiovascular disease risk(14,15). Advances in high-throughput proteomics technologies, such as Olink and SomaScan, now enable simultaneous quantification of thousands of circulating proteins in population-scale cohorts(16). Previous studies have demonstrated that proteomic signatures can improve cardiovascular risk prediction beyond clinical risk factors in both primary(17–19) and secondary prevention populations(20–22). However, most proteomics investigations have focused on incident (primary prevention) cardiovascular disease, while studies in secondary prevention have typically been limited by small sample sizes, low ethnic diversity, and lack of assessment in prospective cohorts. As a result, the incremental value and generalisability of proteomic risk stratification for recurrent events remain uncertain. Further, with the implementation of population-scale proteomics for primary prevention unlikely, its assessment in focused use cases, such as risk stratification for secondary prevention, may yet yield utility and future implementation.

In this study, we sought to determine whether proteomic profiles can improve prediction of recurrent ASCVD beyond conventional clinical risk scores in multiple ethnic groups. Using plasma measurements of ∼2,900 proteins from ∼9,300 participants with established ASCVD in UK Biobank, we trained proteomic predictors using a regularised machine-learning framework and assessed their performance in six withheld test sets differing by ethnicity and geography. To support potential clinical translation, we further derived a parsimonious set of five proteins that retained predictive performance and are measurable in human blood using commercially available and clinically validated immunoassays.

## Methods

### Study design and populations

This study was conducted within the UK Biobank (UKB), a large prospective cohort of ∼500,000 participants aged 40–69 years at baseline, recruited between 2006 and 2010(23). Participants attended one of 22 assessment centres in England, Wales, and Scotland in the baseline assessment, where they provided detailed information on sociodemographic characteristics, lifestyle behaviours, and disease and medication history; underwent physical, anthropometric, and medical imaging measurements; and donated biological samples(24). Longitudinal follow-up is ongoing through linkage to online questionnaires, national registries, hospital records, and general practice data (https://biobank.ndph.ox.ac.uk/showcase/).

For the present analysis, we included participants who had a history of atherosclerotic cardiovascular disease (ASCVD) prior to baseline and for whom plasma proteomic measurements were available from the UK Biobank Pharma Proteomics Project (UKB-PPP) Phase 2 release (October 2023) (16,25). Ethnicity was defined based on self-reported ethnic background (UK Biobank Field 21000). The present analysis included participants categorized as White British, White Irish, other White background, Black or Black British, or Asian or Asian British; participants reporting any other ethnic background were excluded because of small sample sizes. In total, 9,356 participants met the primary inclusion criteria. Participants whose genetically inferred sex did not match recorded sex and those with more than one self-reported ethnic background recorded across study records were excluded as a quality-control step.

The UK Biobank study received ethical approval from the North West Multi-Centre Research Ethics Committee (REC reference 11/NW/03820, 16/NW/0274, 21/NW/0157). All participants provided written informed consent. Analyses for this study were conducted under application number 608471.

### Plasma proteomic profiling

Baseline EDTA plasma samples from Phase 1 of the UK Biobank Pharma Proteomics Project (UKB-PPP) were analysed to capture 2,923 unique proteins across 53,064 participants using the Olink® Explore 3072 Proximity Extension Assay (PEA). Phase 1 includes a large randomly selected baseline subset together with additional consortium-selected and COVID-19 repeat-imaging participants(25). Aliquots stored at −80°C in the UK Biobank automated archive were shipped on dry ice to Olink (Uppsala, Sweden) for proteomic quantification. Sample handling, assay procedures, quality control and preprocessing protocols have been described previously (25). Briefly, Olink’s internal run- and sample-level QC procedures incorporated incubation, extension, and amplification controls for all samples, while external negative, pooled plate, and pooled sample controls were used to define limits of detection, harmonize plates, and monitor assay precision. Normalized Protein eXpression values (log-2 scale) were derived after control-based normalization and within- and between-batch normalization. Outliers, observations with QC or assay warnings, and likely sample swaps were excluded. The resulting dataset showed low intra-individual variability and minimal batch or plate effects(25). For the present analysis, four proteins with missing values in more than 25% of samples and 142 participants with more than 70% of missing protein measurements were excluded from analysis. Missing protein values were imputed using the mean of observed values(26,27). The remaining protein levels were normalized using rank-based inverse normal transformation.

### Clinical risk factors

Baseline clinical characteristics and risk factors were derived from the UK Biobank assessment visit, linked electronic health records, and questionnaire data. The primary set of clinical variables corresponded to the predictors included in the SMART and SMART2 risk scores for secondary cardiovascular prevention(11,28). These included baseline age, self-reported sex, current smoking status (yes/no), systolic blood pressure (mmHg), non-HDL cholesterol (mmol/L), high-sensitivity C-reactive protein (hsCRP; mg/L), and renal function as estimated glomerular filtration rate (eGFR; mL/min/1.73 m²) using the CKD-EPI equation(29), presence of diabetes, coronary artery disease, cerebrovascular disease, abdominal aortic aneurysm, peripheral arterial disease, years since first ASCVD diagnosis, and baseline antithrombotic drug use. Baseline use of aspirin or equivalent therapy (including other antiplatelet or oral anticoagulant drugs) was derived from interview data and primary care records. Missing baseline covariate values were handled using single stochastic imputation by predictive mean matching from a baseline clinical imputation model including age, sex, current smoking, systolic blood pressure, non-HDL cholesterol, C-reactive protein, serum creatinine, antithrombotic use, and the auxiliary variables BMI and HDL cholesterol (30). Imputation was performed in the full available cohort, and analyses were subsequently restricted to participants with Olink measurements.

### Outcomes of Interest

Both prior and recurrent cardiovascular events were defined using the International Classification of Diseases, 10th Revision (ICD-10), and Operating Procedure Codes Supplement (OPCS-4) procedural codes. Disease definitions were based on UK Biobank hospital episode statistics (HES) and death registries. A full list of ICD-10 and OPCS-4 codes used to define each endpoint are provided in **Supplementary Table 1**.

Prior ASCVD at baseline was defined in accordance with established guidelines and included history of acute coronary syndrome, myocardial infarction, stable or unstable angina or coronary or other arterial revascularization, stroke, transient ischemic attack, and peripheral artery disease including aortic aneurysm(31). We additionally considered the presence of cardiac and vascular implants and grafts, as well as coronary revascularization procedures, including coronary artery bypass grafting and percutaneous coronary interventions. Events occurring within 30 days before the baseline assessment were excluded to ensure stable disease status.

The primary outcome was recurrent ASCVD, defined as the first new event occurring more than 30 days after baseline among participants with prior ASCVD. Endpoint definitions followed those of the SMART2 risk score(11) and included non-fatal myocardial infarction, non-fatal stroke, and fatal cardiovascular disease. Conditions excluded from the definition of recurrent events were myocarditis, subarachnoid haemorrhage, subdural haemorrhage, cerebral aneurysm, cerebral arteritis, and moyamoya disease. In total, 1,378 participants experienced a recurrent ASCVD event. As a secondary outcome, we assessed an expanded composite of incident major adverse cardiovascular events (MACE), defined as acute coronary syndrome, acute ischemic stroke, hospitalization for heart failure, cardiac arrest, coronary revascularization procedures, or cardiovascular death, plus transient ischemic attack (TIA) occurring more than 30 days after baseline. In total, 5,222 participants experienced a recurrent MACE event.

### Statistical and machine learning analyses

#### Proteomic feature selection

Participants were partitioned into a discovery cohort for feature selection and model training (training dataset), and six validation cohorts (testing datasets) for testing across multiple ethnic and geographic groups. The training dataset comprised a randomly sampled 70% of White British participants (n=5,467) recruited from assessment centres in England and Wales. The testing datasets (total n=3,889), which were entirely excluded from model development and used only for model evaluation, comprised the held-out 30% of White British participants from England and Wales (n=2,342), White British participants from Scotland (n=605), and participants of White Irish (n=294), Other White background (n=251), Asian or Asian British (n=197), and Black or Black British (n=200) ethnicities.

To identify a robust set of predictive protein biomarkers for recurrent cardiovascular events, we applied a machine learning-based feature selection framework using elastic-net regularization (a combination of L1 and L2 penalties)(32,33) on the 2,919 plasma proteins, optimizing hyperparameters and mitigating overfitting through 5-fold cross-validation repeated 10 times within the training dataset. This approach generated sparse, interpretable models(32) in high-dimensional proteomic data where proteins are correlated and yielded an initial set of predictive proteins associated with recurrent cardiovascular events. Model discrimination was evaluated using area under the receiver operating characteristic curve (AUC).

#### Derivation and validation of proteomic prediction models

Proteins selected by elastic-net were fit in Cox proportional hazards models to derive proteomic prediction models for recurrent events in the training dataset. For protein-based models, coefficients were estimated in the training dataset and then applied unchanged to the predefined independent testing datasets of participants from different ethnic and geographic backgrounds, with the time to first recurrent event as the outcome and years since baseline sampling as the time scale. The SMART2 score was calculated in each dataset as a linear predictor using the published coefficients from the SMART2 study(11). We first compared the proteomic score with SMART2, and then assessed whether predictive performance was improved by additionally incorporating age and sex or by combining the proteomic score with the SMART2 score. For each model, analyses included only participants with non-missing outcome data and all predictors required for that model.

To derive a parsimonious set of proteins suitable for potential translation into a targeted, clinically scalable test, we selected a five-protein set with commercially available human ELISA kits with the largest absolute coefficient weights from the elastic-net model. A Cox model including these five proteins was then fitted in the training dataset and evaluated in the predefined testing datasets using the same validation framework as for the full protein model. Its performance was compared with SMART2 and with the corresponding combined model.

#### Model performance assessment

The performance of Cox models was quantified using Harrell’s C-index. Calibration was assessed at a fixed 10-year time horizon. For SMART2, the published linear predictor was computed from the model coefficients and converted to 10-year risk using the published prediction algorithm for the European low-risk region. For the locally fitted Cox models, 10-year predicted risks were derived from the model linear predictor and the baseline cumulative hazard at 10 years. Grouped calibration plots were constructed by dividing participants into deciles of predicted 10-year risk and plotting the mean predicted risk against the observed 10-year risk estimated using Kaplan–Meier methods, with 95% confidence intervals for the observed risk. Global calibration was summarized by comparing the mean predicted 10-year risk with the observed 10-year risk and by calculating the observed-to-expected (O/E) ratio.

### Data and Code Availability

All data described are available through UK Biobank subject to approval from the UK Biobank access committee. See https://www.ukbiobank.ac.uk/enable-your-research/apply-for-access for further details. Code underlying this paper is available upon request.

## Results

The study included 9,356 participants in the UK Biobank with established ASCVD at baseline and available plasma proteomic profiles through the UKB-PPP (**Figure 1**). The cohort comprised 45% females, with a median age of 62.2 years (interquartile range (IQR) 56.1–66.3) at baseline assessment. During a median follow-up period of 13.5 years (IQR 12.7–14.4), 1,378 participants (14.7%) experienced at least one recurrent ASCVD event, while 5,222 (55.8%) experienced at least one major adverse cardiovascular event (MACE). Participant characteristics across predefined training and validation sets, including ethnic and geographic strata are summarized in **Table 1** and **Supplementary Table 2**.

**Figure 1.**
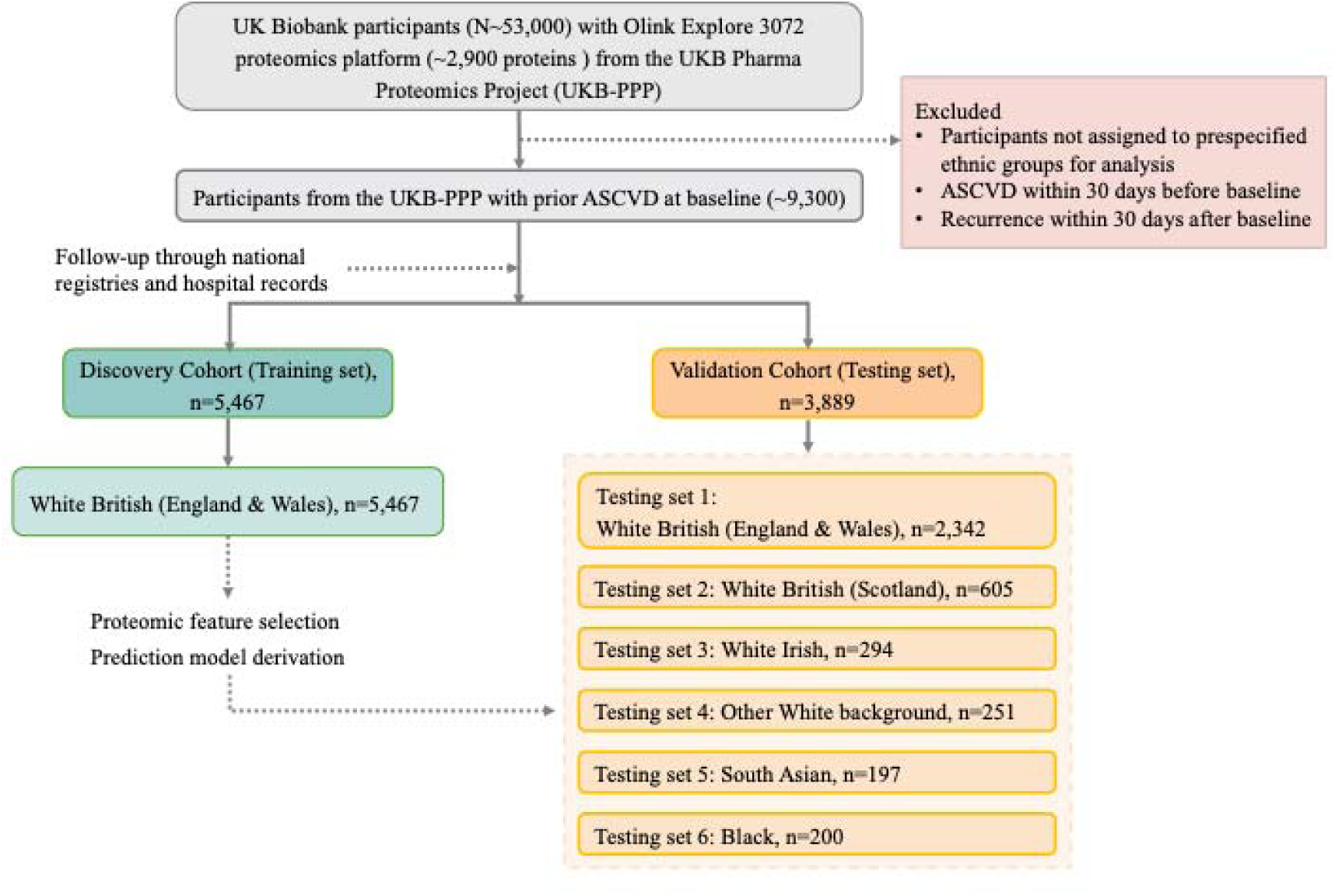
Overview of study design and workflow.

**Table 1.** Characteristics of UK Biobank Pharma Proteomics Project participants included in the present analysis of recurrent ASCVD (n = 9,356). Values are presented as median (interquartile range) for continuous variables and n (%) for categorical variables.

|  | Discovery cohort for training | Validation cohorts for testing |  |  |  |  |  |
| --- | --- | --- | --- | --- | --- | --- | --- |
| Dataset | White British (England & Wales) | White British (England & Wales) | White British (Scotland) | White Irish | Other White Background | Asian or Asian British | Black or Black British |
| N | 5467 | 2342 | 605 | 294 | 251 | 197 | 200 |
| Length of follow-up, years | 13.50 (12.69–14.26) | 13.47 (12.62–14.27) | 14.82 (14.27–15.08) | 13.41 (12.56–14.35) | 13.42 (12.70–14.16) | 13.15 (12.57–13.90) | 13.12 (12.63–13.81) |
| Recurrent ASCVD, n (%) | 810 (14.8%) | 347 (14.8%) | 85 (14.0%) | 47 (16.0%) | 33 (13.1%) | 28 (14.2%) | 28 (14.0%) |
| Baseline age, years | 62.42 (56.77–66.31) | 62.46 (56.70–66.31) | 61.51 (55.27–66.43) | 62.37 (56.73–66.69) | 60.04 (53.32–65.22) | 57.61 (51.06–64.66) | 55.17 (49.53–62.78) |
| Female sex, n (%) | 2415 (44.2%) | 1038 (44.3%) | 283 (46.8%) | 140 (47.6%) | 130 (51.8%) | 86 (43.7%) | 118 (59.0%) |
| BMI, kg/m<sup>2</sup> | 27.58 (24.84–30.88) | 27.66 (25.02–30.78) | 27.42 (25.00–30.54) | 27.33 (24.48–30.33) | 27.68 (24.15–30.91) | 27.37 (24.67–30.29) | 28.66 (25.83–31.89) |
| Systolic blood pressure (mmHg) | 138.0 (126.0–150.0) | 138.0 (127.0–151.0) | 139.0 (128.0–152.0) | 134.0 (124.0–146.8) | 133.0 (122.0–146.0) | 134.0 (122.0–146.0) | 137.0 (126.0–148.0) |
| Diastolic blood pressure (mmHg) | 81.0 (74.0–88.0) | 81.0 (74.0–88.0) | 82.0 (75.0–89.0) | 80.0 (72.0–86.0) | 79.0 (73.2–87.0) | 81.0 (74.0–88.0) | 83.0 (76.8–91.0) |
| Total cholesterol (mmol/L) | 5.24 (4.42–6.14) | 5.25 (4.41–6.20) | 5.15 (4.37–6.19) | 5.39 (4.41–6.23) | 5.24 (4.42–6.19) | 4.92 (4.03–5.60) | 5.02 (4.17–5.82) |
| HDL cholesterol (mmol/L) | 1.31 (1.09–1.56) | 1.31 (1.09–1.56) | 1.29 (1.06–1.58) | 1.36 (1.12–1.66) | 1.37 (1.16–1.61) | 1.16 (1.00–1.46) | 1.35 (1.15–1.64) |
| LDL cholesterol (mmol/L) | 3.21 (2.63–3.92) | 3.23 (2.63–3.95) | 3.18 (2.57–4.01) | 3.28 (2.51–3.85) | 3.22 (2.63–3.92) | 3.03 (2.36–3.66) | 3.04 (2.46–3.62) |
| Triglycerides (mmol/L) | 1.61 (1.14–2.29) | 1.61 (1.15–2.30) | 1.63 (1.13–2.42) | 1.51 (1.08–2.19) | 1.47 (1.09–2.17) | 1.66 (1.17–2.32) | 1.08 (0.80–1.47) |
| C-reactive protein (mg/L) | 1.66 (0.79–3.47) | 1.67 (0.81–3.47) | 1.76 (0.79–3.83) | 1.92 (0.85–4.20) | 1.38 (0.66–2.85) | 1.89 (0.84–3.39) | 1.69 (0.71–3.23) |
| Creatinine (μmol/L) | 73.9 (63.6–85.5) | 74.2 (63.8–86.0) | 72.1 (61.3–83.2) | 73.5 (62.3–83.3) | 71.8 (62.7–84.1) | 69.8 (60.3–80.5) | 77.1 (67.1–89.3) |
| Current smoking, n (%) | 596 (10.9%) | 249 (10.6%) | 93 (15.4%) | 63 (21.4%) | 32 (12.7%) | 18 (9.1%) | 23 (11.5%) |
| Diabetes mellitus, n (%) | 534 (9.8%) | 253 (10.8%) | 58 (9.6%) | 24 (8.2%) | 38 (15.1%) | 60 (30.5%) | 35 (17.5%) |
| Antithrombotic medication, n (%) | 2162 (39.5%) | 916 (39.1%) | 276 (45.6%) | 114 (38.8%) | 99 (39.4%) | 93 (47.2%) | 51 (25.5%) |

### Derivation and performance of proteomic predictors for recurrent ASCVD

To identify proteomic signatures predictive of recurrent ASCVD, we employed a regularized machine learning approach that simultaneously performs feature selection and addresses potential multi-collinearity across 2,919 plasma proteins captured by the Olink® Explore 3072 PEA. The model development utilized a training set comprising 70% of randomly selected participants of White British ethnicity from England and Wales (n=5,467; **Figure 1**). Through repeated 5-fold cross-validation, the best-performing model retained 38 proteins (**Supplementary Table 3**) with a mean cross-validated AUC of 0.723 in the training dataset.

To assess generalizability and cross-ethnicity transferability, we evaluated the proteomic predictors in six prespecified testing datasets defined by self-reported ethnicity and geography. These included held-out White British participants from England and Wales, White British participants from Scotland, and participants of White Irish, Other White, Asian or Asian British, and Black or Black British ethnicities. The prediction performance for recurrent ASCVD was highest in White Irish (**Figure 2**; AUC: 0.815, 95% Confidence Interval: 0.753–0.875) and Other White ethnicity (AUC: 0.810, 95% CI: 0.742–0.872), and remained robust in White British from England & Wales (AUC: 0.720, 95% CI: 0.693–0.748) as well as Scotland (AUC: 0.723, 95% CI: 0.668–0.775). However, the proteomic predictor was somewhat attenuated in Asian or Asian British (AUC: 0.671, 95% CI: 0.564–0.771) and Black ethnicities (AUC: 0.645, 95% CI: 0.552–0.732).

**Figure 2.**
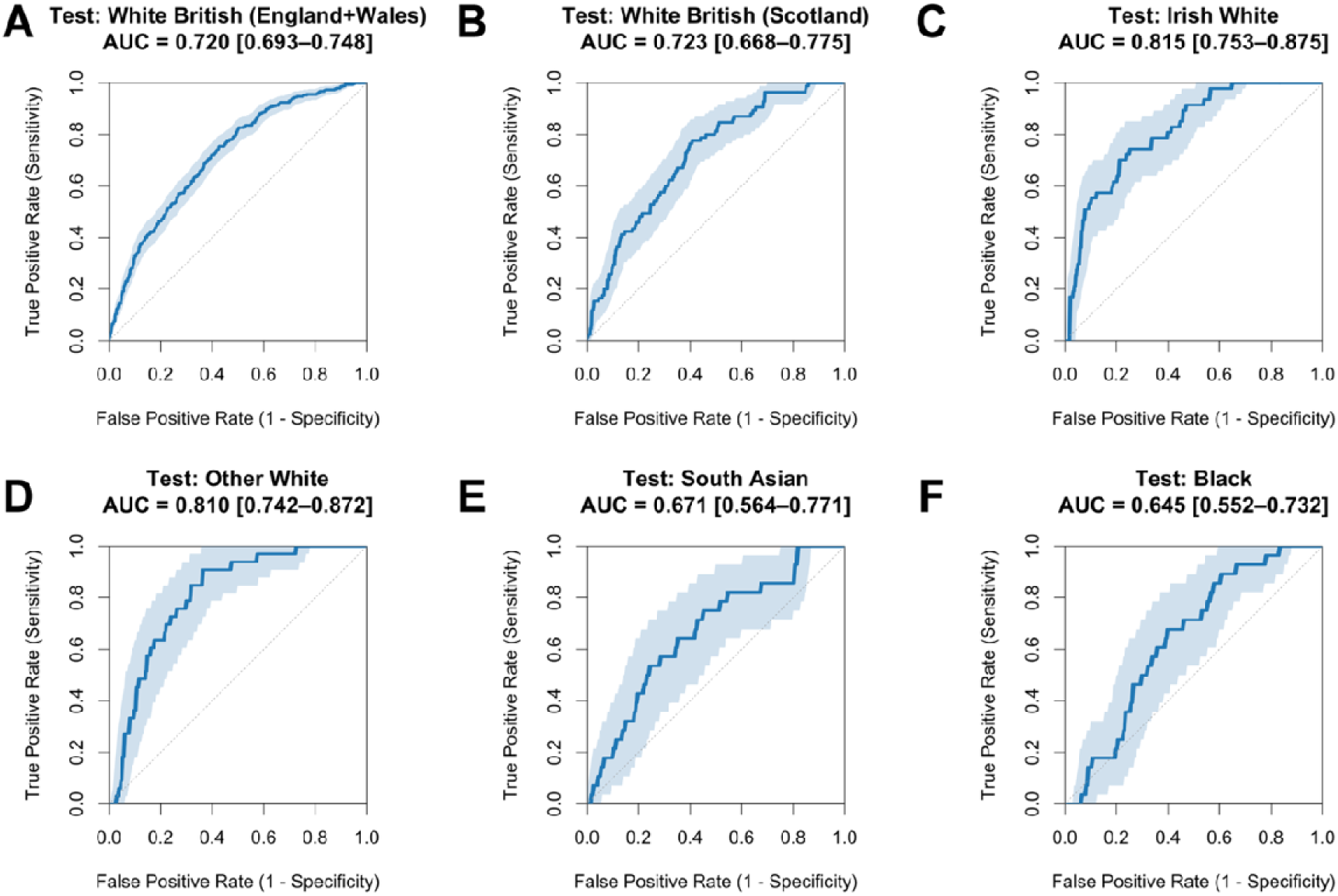
Performance of machine-learning feature selection for proteomic predictors of recurrent ASCVD across independent testing sets. The ROC curves with 95% confidence bands for the proteomic model derived using 2,919 plasma proteins in White British (England & Wales) training sets and assessed in six independent testing sets for **(A)** White British (England & Wales, *n* = 2,342; 347 events), **(B)** White British (Scotland, *n* = 605; 85 events), **(C)** White Irish (*n* = 294; 47 events), **(D)** Other White background (*n* = 251; 33 events), **(E)** Asian or Asian British (*n* = 197; 28 events), and **(F)** Black or Black British (*n* = 200; 28 events) ethnicities.

When the same feature selection procedure was applied to the MACE endpoint, 160 proteins (**Supplementary Table 4**) were selected by the best-performing model and yielded a cross-validated AUC of 0.663, likely reflecting the greater aetiological heterogeneity of this composite endpoint. In the testing sets, proteomic predictors for MACE exhibited lower but more consistent performance across ethnic and geographic groups as compared to ASCVD, with AUCs ranging from 0.632 (95% CI: 0.555–0.710) in in individuals of Black or Black British ethnicity to 0.682 (95% CI: 0.608–0.756) in those of Asian or Asian British ethnicity (**Supplementary Figure 1**).

### Incremental improvements beyond SMART2 clinical risk score

To evaluate whether proteomic profiling provides incremental predictive value beyond established clinical risk assessment, we compared a protein score derived from pre-selected proteins, the SMART2 risk score(11), and their combination. To derive the protein score, we fit a Cox proportional hazards model incorporating the 38 pre-selected proteins using the same training set (i.e. 70% of White British participants from England and Wales) and applied the derived coefficients unchanged to the six independent testing sets to compare model performance across different ethnic and geographic backgrounds.

The protein score consistently outperformed SMART2 for predicting recurrent ASCVD across all testing sets (**Figure 3**). Across the six testing groups, the protein score yielded a mean C-index of 0.743, ranging from 0.654 (95% CI, 0.554–0.747) in Black or Black British participants to 0.826 (95% CI, 0.773–0.879) in White Irish participants. By comparison, SMART2 achieved a mean C-index of 0.653 across the six testing groups, ranging from 0.590 (95% CI, 0.470–0.709) in Black or Black British participants to 0.727 (95% CI, 0.629–0.820) in participants of Other White background.

**Figure 3.**
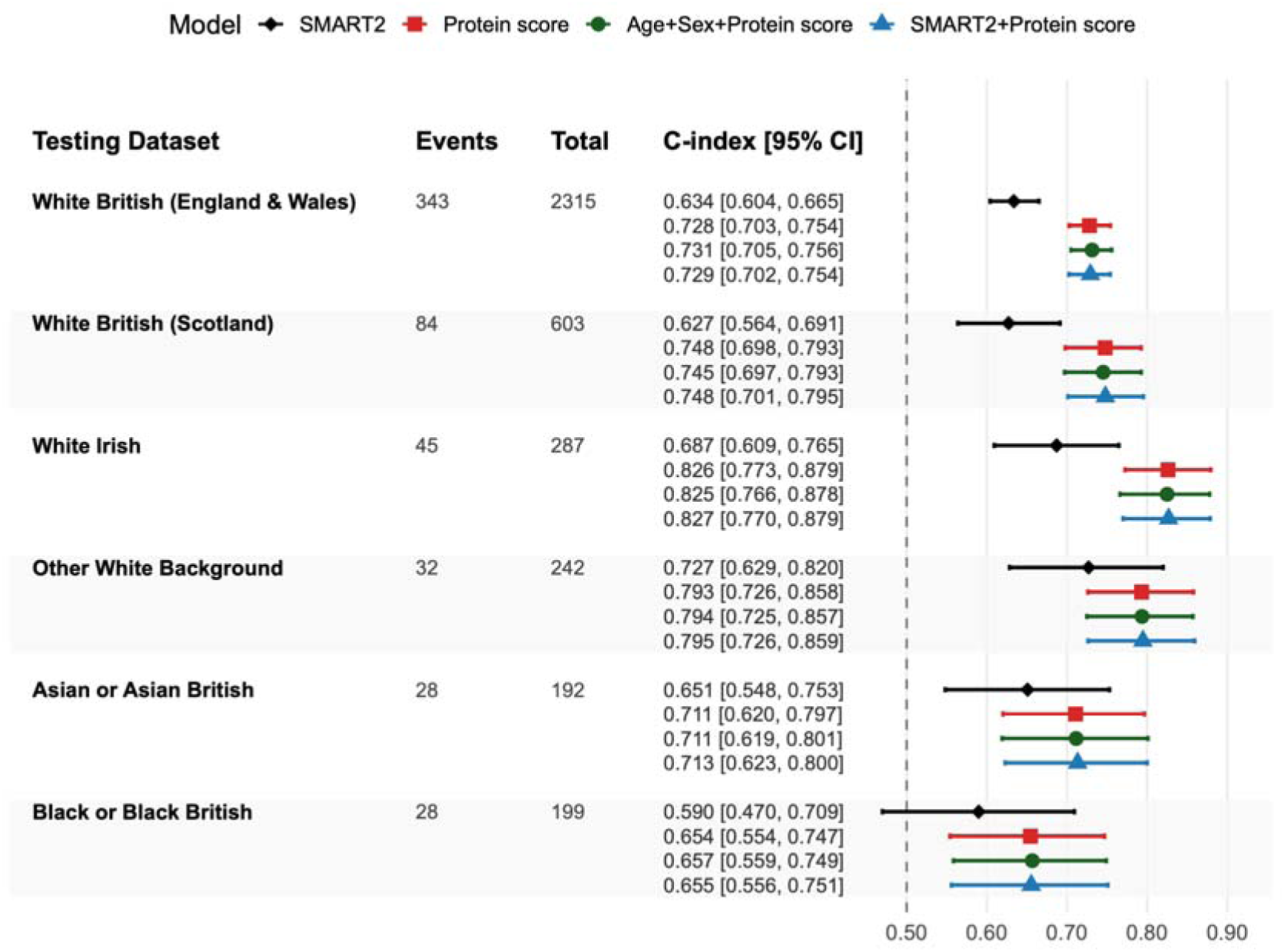
Prediction performance of protein, clinical, and combined models for recurrent ASCVD across ethnic and geographic subgroups. Forest plot of Harrell’s C-index with 95% confidence intervals for SMART2, the protein score, a model of age + sex + protein score, and the combined model integrating SMART2 and the protein score in each testing dataset.

Combining the protein score with the SMART2 risk score yielded substantial improvements in ASCVD prediction compared with SMART2 alone (**Supplementary Figure 2**). Significant gains were observed in White British participants from England and Wales, White British participants from Scotland, and White Irish participants, with the largest ΔC-index observed in White Irish participants (0.140, 95% CI 0.074–0.205; P<0.001). While statistical power was limited due to smaller sample sizes and event numbers, the combined proteomic and SMART2 predictor showed a similar trend in Asian or Asian British participants (ΔC-index 0.062, 95% CI −0.056 to 0.184; P=0.297) and in Black or Black British participants (ΔC-index 0.066, 95% CI −0.071 to 0.208; P=0.385). Notably, combining SMART2 with the protein score yielded performance similar to that of the protein score alone (**Figure 3**) with ΔC-indices ranging from 0.000 to 0.003 across testing groups (**Supplementary Figure 2**). A model combining the protein score with age and sex provided nearly identical performance to the protein score alone across all testing sets (**Figure 3**).

For prediction of MACE, the model of 160 selected proteins demonstrated moderate performance (mean C-index 0.610) and consistently outperformed the SMART2 risk score (mean C-index 0.575) across all testing sets (**Supplementary Figure 3**). Combining the protein score for MACE with SMART2 produced modest but statistically significant improvements over SMART2 alone in White British participants from England, Wales and Scotland, whereas gains over the proteomic model alone were minimal overall (**Supplementary Figure 4**).

### A parsimonious protein panel with commercially available clinical assays

Proteomics technologies are generally not available in clinical practice; however, clinical laboratories have ready access to Enzyme-Linked Immunosorbent Assays (ELISAs) for clinical testing. To facilitate potential clinical translation, we derived a parsimonious set of proteins that preserved the majority of predictive capacity. From the 38 proteins which made up the full protein score for ASCVD, the five proteins with the largest weights in the model and with commercially available human ELISA assays were selected. The 5-protein set was fit in the training set using a Cox model and evaluated for predictive performance across the six independent testing sets.

The 5-protein set for recurrent ASCVD comprised growth differentiation factor 15 (GDF15), N-terminal pro-B-type natriuretic peptide (NT-proBNP), WAP four-disulfide core domain protein 2 (WFDC2), renin (REN), and matrix metallopeptidase 12 (MMP12). The 5-protein score maintained robust predictive performance for recurrent ASCVD across testing sets (mean C-index 0.718; Figure 4), with C-index ranging from 0.621 (95% CI, 0.531–0.708) in Black or Black British participants to 0.818 (95% CI, 0.743–0.880) in participants of Other White background. Consistent with findings for the full protein score, the 5-protein score showed higher predictive performance than SMART2 overall, except Asian or Asian British participants (**Figure 4**). Combining the 5-protein score with SMART2 improved performance compared with SMART2 alone in White British participants from England and Wales (ΔC-index 0.084, 95% CI 0.054–0.112; P<0.001; **Supplementary Figure 5**), White British participants from Scotland (ΔC-index 0.082, 95% CI 0.016–0.143; P=0.014), and White Irish participants (ΔC-index 0.115, 95% CI 0.046–0.182; P=0.001), with borderline evidence in participants of Other White background (ΔC-index 0.093, 95% CI −0.000 to 0.192; P=0.051). No clear improvement was observed in Asian or Asian British participants (ΔC-index 0.000, 95% CI −0.125 to 0.128; P=1.000) or Black or Black British participants (ΔC-index 0.039, 95% CI −0.082 to 0.164; P=0.543). In contrast, the model combining SMART2 and the 5-protein score yielded only minimal additional gains over the 5-protein score alone, with ΔC-indices ranging from −0.001 to 0.008 across testing datasets. A model combining age, sex and the 5-protein score showed only modestly higher performance than the 5-protein score alone (**Figure 4**). A 5-protein score for the MACE outcome resulted in similar trends (**Supplementary Figure 6&7**).

**Figure 4.**
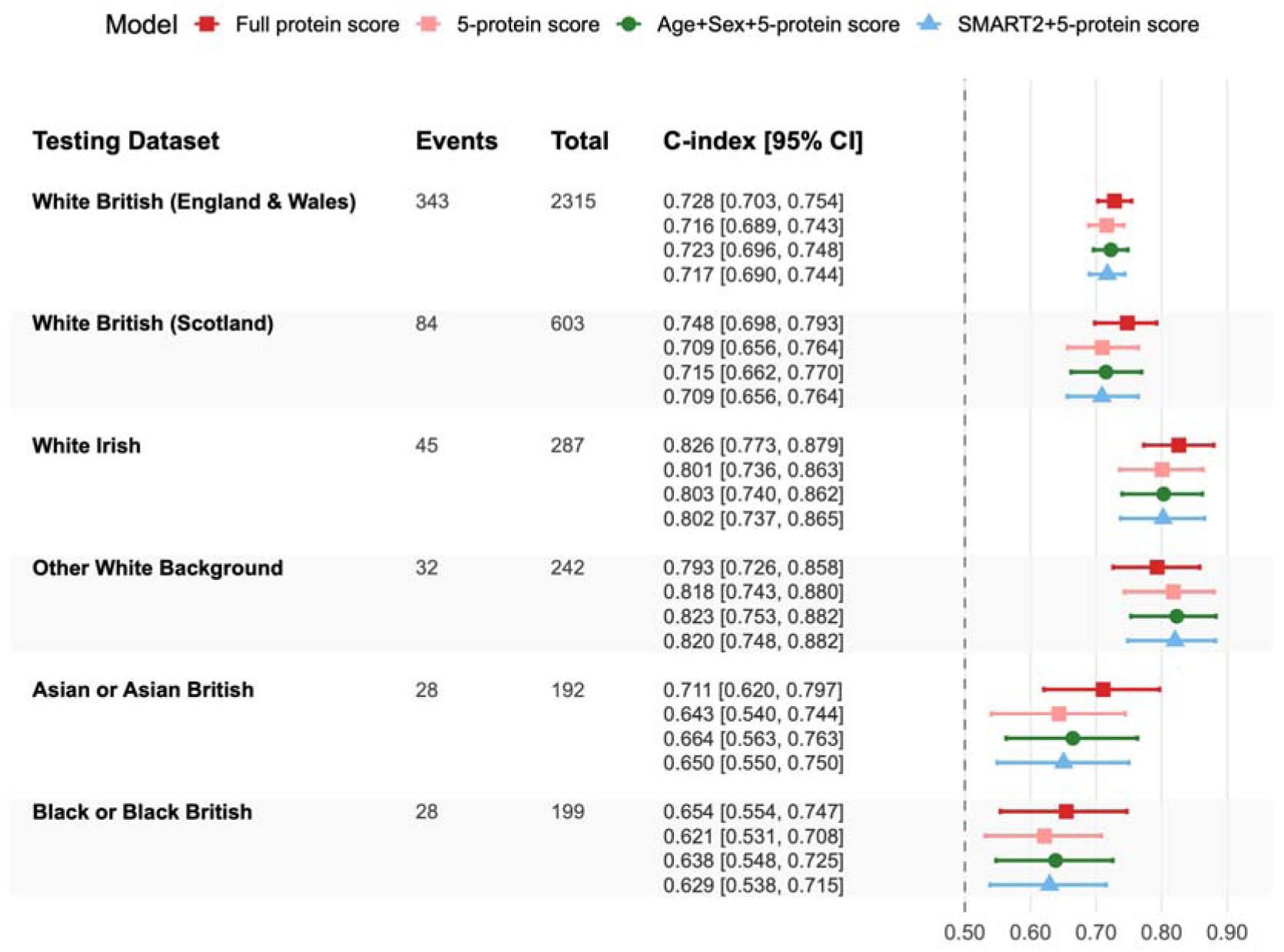
Comparative performance of the full protein score and the 5-protein score for recurrent ASCVD. Forest plot of Harrell’s C-index with 95% confidence intervals for the full protein score, the 5-protein score, the age+sex+5-protein score, and the combined model integrating SMART2 and the 5-protein score in each testing dataset.

### Calibration and risk stratification

The full protein score and the 5-protein score showed reasonable agreement between predicted and observed 10-year recurrent ASCVD risk in most testing datasets (**Supplementary Figure 8**). In White British participants from England & Wales and Scotland, O/E ratios for the full protein score and the 5-protein score ranged from 0.997 to 1.042.

Kaplan-Meier survival curves for recurrent ASCVD showed clear risk stratification across quintiles of both the full protein score and the 5-protein score in the White British (England & Wales) testing set (**Figure 5**). The 5-protein score preserved much of the risk stratification seen with the full protein score, when assessing the 10-year cumulative event probabilities in the highest (approx. 27.4% vs 29.0%), middle (9.6% vs 8.5%) and lowest (2.4% vs 2.2%) quintiles, respectively. Similar patterns were observed in White British participants from Scotland.

**Figure 5.**
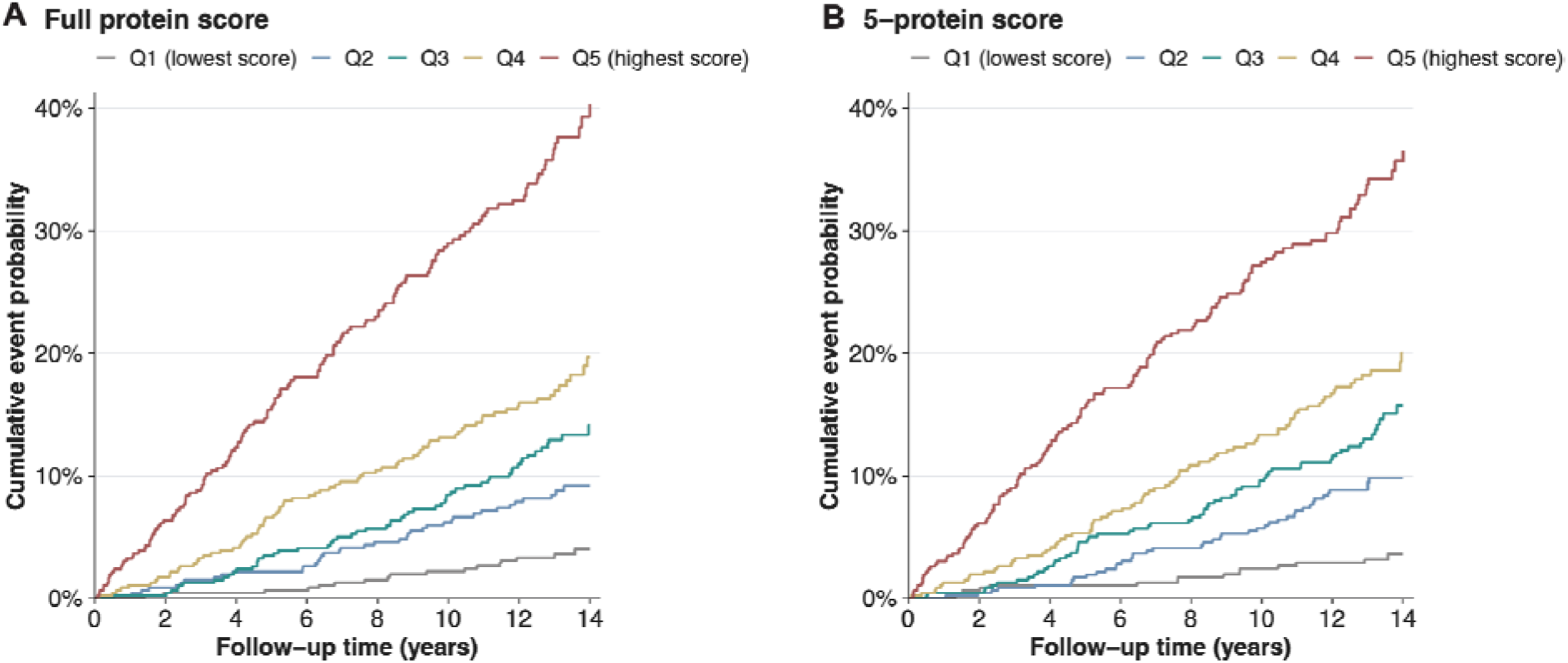
Recurrent ASCVD-free survival stratified by quintiles of protein score. Kaplan–Meier curves are shown for (A) the full protein score and (B) the 5-protein score in the White British (England & Wales) testing set, plotted as cumulative event probability (1 – recurrent ASCVD-free survival). Participants were grouped into quintiles within the testing set according to the distribution of each score, with Q1 indicating the lowest score quintile and Q5 the highest.

## Discussion

Accurate risk stratification after an initial ASCVD event is central to secondary prevention, particularly for guiding treatment intensification and follow-up strategies. Our study demonstrates that plasma proteomics substantially improves prediction of recurrent ASCVD beyond conventional clinical risk factors. In a large, population-scale cohort, the proteomic models consistently outperformed a guideline-recommended clinical risk score for recurrent ASCVD (SMART2) across multiple ethnic and geographic groups, demonstrating transportability of the protein-based signal for recurrent ASCVD prediction. Adding proteomic predictors to SMART2 significantly improved prediction performance over SMART2 alone, but did not improve prediction beyond the protein-based predictor alone, indicating that circulating proteins both captured information provided by conventional risk factors as well as contributed additional complementary prognostic information. A parsimonious 5-protein score measurable with commercially available human ELISAs retained almost all the predictive signal of the full protein score, suggesting that wide-angle proteomic profiling technologies may not be necessary for clinical implementation but are useful for identifying the most relevant easily quantified predictive biomarkers. Taken together, these results support a plasma protein score as a promising complement to clinical risk prediction tools for secondary prevention of cardiovascular disease.

Despite advances in acute and chronic management, patients with established ASCVD remain at persistently high risk of recurrent events, with substantial heterogeneity between individual (10,12). While most cardiovascular risk prediction models target primary prevention of a first event (e.g., SCORE2(34), QRISK3(35), the Pooled Cohort Equations(7), and the Framingham Risk Score(36)), fewer tools address the critical need for risk stratification in secondary prevention. Available secondary-prevention models for recurrent cardiovascular events include the SMART2 risk score for recurrent ASCVD(11), TRS2P score for risk stratification in patients undergoing coronary angiography(37), and SMART-REACH risk score for estimation of life expectancy(13). Despite multiple clinical predictors, discriminative accuracy remains moderate, indicating a plateau of prediction performance for models constrained to conventional risk factors (likely due to index event bias), and highlights the need for more biologically proximal markers that reflect active disease processes. Our findings indicate that large-scale proteomic profiling may help overcome this limitation by integrating signals from diverse pathophysiological pathways underlying recurrent risk.

The proteins comprising the proteomic predictor of recurrent ASCVD risk span multiple pathophysiological domains, including cardiac stress (e.g. NT-proBNP, NPPB)(38–40), cellular stress and inflammation (e.g. GDF15(41,42)), innate immune modulation (e.g. WFDC2(43)), extracellular matrix (ECM) remodelling (e.g. MMPs(44)), endothelial angiogenic signalling (e.g. VEGFA(45)), lipid metabolism (e.g. ANGPTL3(46), APOC1(47), FABP1(48)), renal injury (e.g. HAVCR1(49)), and neuronal injury(e.g. NEFL(50)). These domains are broadly consistent with prior proteomic prediction studies of cardiovascular outcomes, although direct comparisons should be interpreted cautiously because published models differed in clinical setting, composite endpoint, follow-up horizon, and proteomic platform. Notably, MMP12 was retained both in the 9-protein score using aptamer-based proteomic profiling by Ganz et al.(20) for 4-year risk of myocardial infarction, stroke, heart failure, and all-cause death in stable coronary heart disease and the 27-protein model reported by Williams et al.(14) using the SomaScan platform for 4-year risk of myocardial infarction, stroke, heart failure, or death; NT-proBNP also overlapped with Williams et al.(14); in line with this, the Olink-based secondary-prevention study by Nurmohamed et al. identified NT-proBNP and GDF-15 among its most informative predictors(22). Among the 5-protein panel for recurrent ASCVD, NT-proBNP is an established prognostic biomarker and a guideline-recommended marker for heart failure risk stratification, whereas GDF-15 is a well-established prognostic biomarker of cardiovascular risk (51–53). Circulating MMP12 has been associated with greater atherosclerotic burden and incident coronary events, and converging genetic and experimental data further implicate MMP12 in plaque remodelling and large-artery atherosclerotic stroke (54,55). Circulating renin (REN) reflects RAAS (renin-angiotensin-aldosterone system) activation, and higher plasma renin activity has been associated with myocardial infarction and cardiovascular mortality(56,57). WFDC2 (HE4), one of the strongest predictors, has received limited attention in cardiovascular research despite emerging evidence linking it to fibrosis, heart failure and renal dysfunction(58–62). Originally characterised as a secreted WAP-domain protein implicated in epithelial innate immunity and host defence(63,64), WFDC2’s putative immunomodulatory and protease-regulatory functions may provide a plausible biological link between inflammation, ECM remodelling, and recurrent ASCVD risk. Taken together, the partial overlap of our selected proteins with prior CVD proteomic scores across SomaScan- and Olink-based studies, alongside clear differences in panel composition, suggests that recurrent ASCVD risk is consistently informed by signals of myocardial stress, matrix remodelling, inflammation, and end-organ injury, while the exact proteins retained in sparse models depend on the clinical phenotype under study and the measurement technology available.

Several limitations warrant consideration. First, we used a single high-throughput proteomics platform (Olink Explore 3072), and cross-platform transferability remains uncertain. Prior studies report modest agreement between protein quantification techniques, with differences in genetic and disease associations that can affect inference(16,65). This limitation should, however, be interpreted in the context of prior large-scale comparisons showing only modest overall concordance between Olink and SomaScan, together with differences in cis-pQTL support and disease associations, indicating that some between-study discrepancies may reflect assay characteristics rather than biology(16). Moreover, not all candidate proteins are represented on all platforms, and even shared targets may differ in analytical performance, which can contribute to discordant cross-study selection(66,67). Notwithstanding this, the recurrence of MMP12 and NT-proBNP across both SomaScan- and Olink-derived cardiovascular scores is reassuring. Replication on an independent platform, particularly one that provides calibrated absolute quantitation, will therefore improve confidence in the translation potential of the 5-protein score to ELISA or other conventional immunoassays. A direct analytical bridging study will be especially important, because although ELISA is likewise antibody-based, it cannot be assumed *a priori* that conventional immunoassays will reproduce the same rank-order discrimination observed with Olink measurements. For NT-proBNP and GDF-15, measurements from earlier Olink PEA panels have shown strong correlation with conventional immunoassays and similar prognostic performance(68). Nevertheless, whether the present five-protein score would retain equivalent prognostic performance after translation to conventional immunoassays requires further investigation. Second, in geographically and ethnically distinct hold-out sets to evaluate generalisability and transferability, performance was attenuated in non-White ethnicities, indicating that larger multi-ethnic cohorts are needed to improve equity of model performance across diverse populations. Third, while we undertook predefined testing across multiple ethnicities, prospective external validation in independent healthcare systems will be essential to establish the broader clinical applicability of these protein-based risk scores. At present, we are not aware of a readily available external cohort with sufficiently large numbers of recurrent ASCVD events, comparable proteomic depth, and suitable clinical phenotyping to provide a close replication of the present analysis. Finally, outcomes were analysed as time from baseline to first recurrent event in our study, and extensions to modelling multiple recurrences and recurrence rates could provide deeper insights into disease dynamics and enable more personalized risk assessment.

In summary, our findings demonstrate that high-throughput proteomics can substantially improve prediction of recurrent ASCVD beyond a guideline-recommended clinical risk score, and that this gain is retained to a large extent in five-protein candidate panel. The proteomic score may offer a potentially generalisable approach to identifying high-risk patients for risk-guided intensification of secondary prevention therapy and closer follow-up across diverse care settings, in line with guideline calls for individualised management(5). By capturing biological signals of active disease beyond conventional clinical predictors, the proteomic score may help identify patients who warrant more intensive secondary-prevention strategies and closer follow-up than would be indicated by clinical risk factors alone. At the same time, clinical translation will require orthogonal assay validation, since it remains to be established whether ELISA or other conventional immunoassays for selected proteins will deliver equivalent prognostic performance in routine care. Future studies should prospectively evaluate, ideally in randomised trials, whether proteomics-guided intensification of secondary-prevention therapy improves clinical outcomes. Incorporating biological markers of active disease alongside clinical variables may support more precise cardiovascular care, with treatment decisions informed by current disease activity as well as population-based risk estimates. Taken together, proteomics may offer a scalable, biology-informed complement to clinical risk scores for prioritising patients at greatest risk of recurrent ASCVD.

## Data Availability

All data described are available through UK Biobank subject to approval from the UK Biobank access committee. See https://www.ukbiobank.ac.uk/enable-your-research/apply-for-access for further details.

## Acknowledgements

This work was supported by core funding from the British Heart Foundation (RG/F/23/110103), NIHR Cambridge Biomedical Research Centre (NIHR203312) [*], BHF Chair Award (CH/12/2/29428), Cambridge BHF Centre of Research Excellence (RE/24/130011), and by Health Data Research UK, which is funded by the UK Medical Research Council, Engineering and Physical Sciences Research Council, Economic and Social Research Council, Department of Health and Social Care (England), Chief Scientist Office of the Scottish Government Health and Social Care Directorates, Health and Social Care Research and Development Division (Welsh Government), Public Health Agency (Northern Ireland), British Heart Foundation and the Wellcome Trust.

Y.L. was supported by the British Heart Foundation and the German Centre for Cardiovascular Research (SP/F/23/150048). S.C.R. was funded by a BHF Cambridge Centre for Research Excellence fellowship (RE/24/130011). S.A.L. was supported by a Canadian Institutes of Health Research postdoctoral fellowship (MFE-171279). A.M.W is supported by the BHF Data Science Centre (HDRUK2023.0239) and as an NIHR Research Professor (NIHR303137). E.D.A. holds a NIHR Senior Investigator Award. M.I. was supported by the UK Economic and Social Research Council (ES/T013192/1), the Lupus Research Alliance (909140), the Munz Chair of Cardiovascular Prediction and Prevention and the NIHR Cambridge Biomedical Research Centre (NIHR203312).

The funders had no role in study design, data collection and analysis, decision to publish, or preparation of the manuscript.

*The views expressed are those of the authors and not necessarily those of the NIHR or the Department of Health and Social Care.

