## Supplementary Figure 1 for "Plasma proteomics improves prediction of recurrent cardiovascular events"

**Supplementary Figure 1. Performance of machine-learning feature selection for proteomic signatures of incident MACE across independent testing datasets.** ROC curves with 95% confidence bands for the protein score derived in White British (England and Wales) training data and applied without refitting to six independent testing datasets: **(A)** White British (England and Wales; *n* = 2,342; 1,321 events), **(B)** White British (Scotland; *n* = 605; 271 events), **(C)** White Irish(*n* = 294; 173 events), **(D)** Other White Background (*n* = 251; 137 events), **(E)** Asian or Asian British (*n* = 197; 129 evets), and **(F)** Black or Black British (*n* = 200; 108 events) participants.

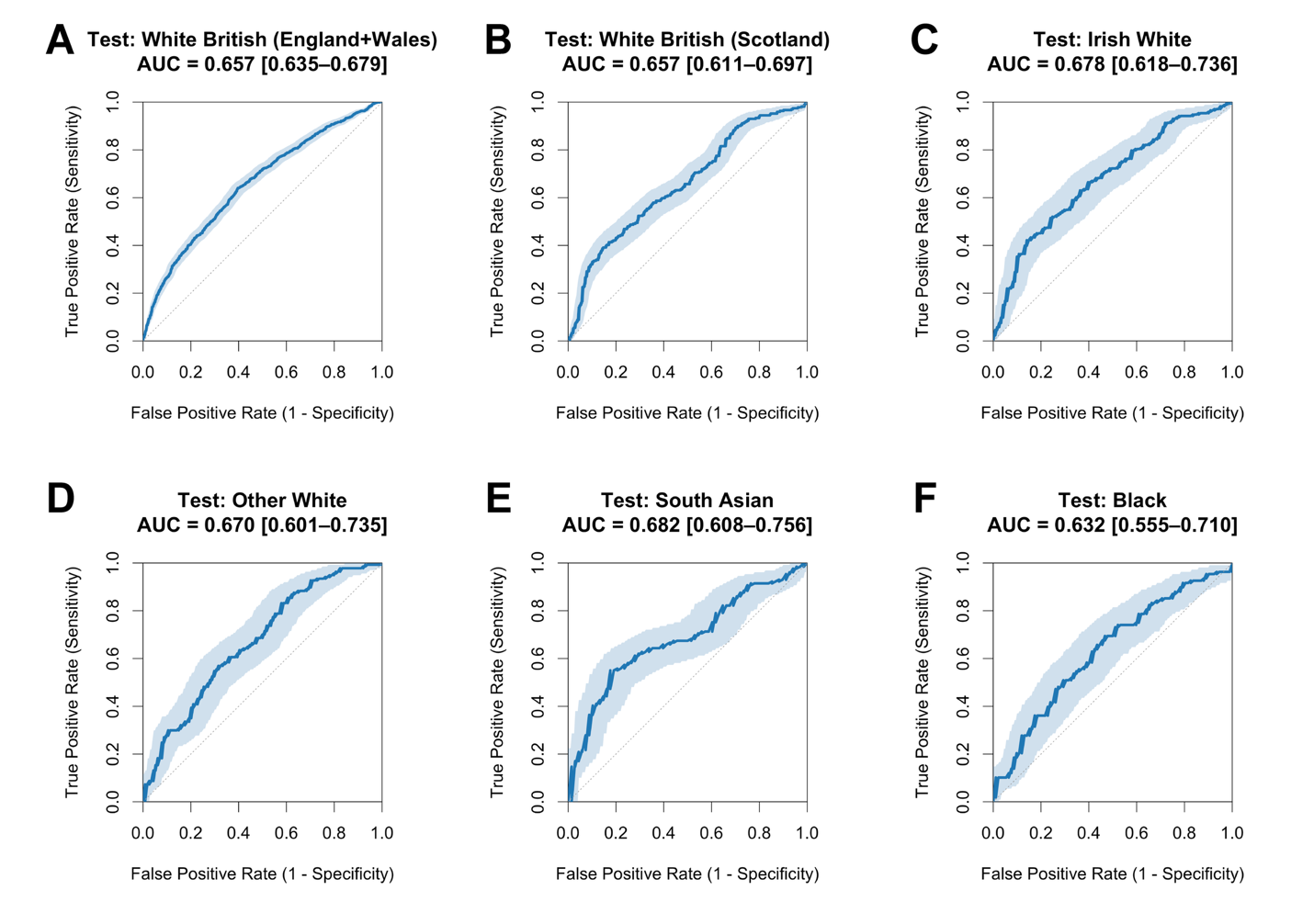
