## Supplementary Figure 3 for "Plasma proteomics improves prediction of recurrent cardiovascular events"

**Supplementary Figure 3. Prediction performance of the protein score, SMART2, and combined models for recurrent MACE across ethnic and geographic subgroups.** Forest plot of Harrell’s C-index with 95% confidence intervals for SMART2, the protein score, a model of age + sex + protein score, and the combined model integrating SMART2 and the protein score in each testing dataset.

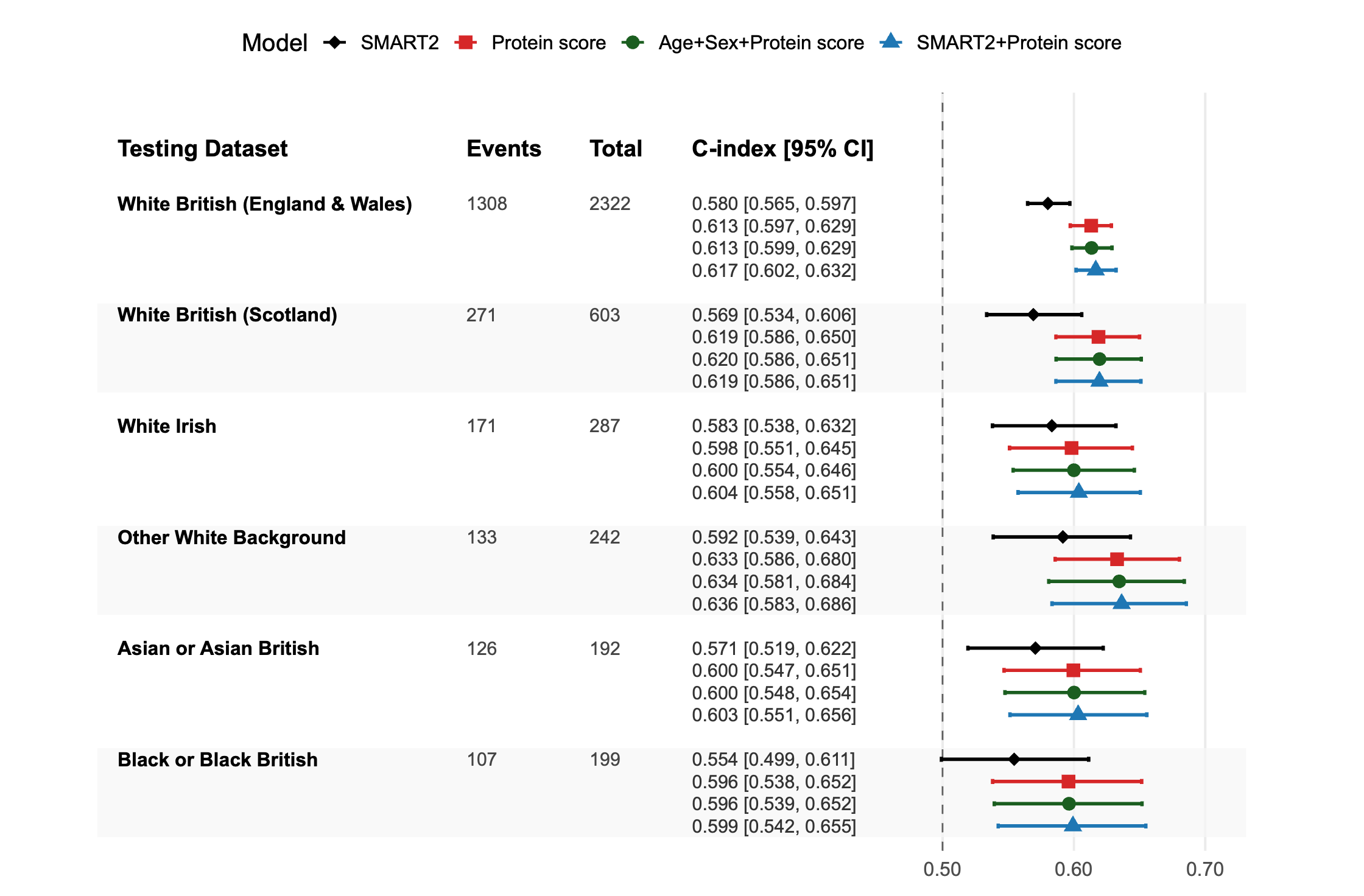
