## Supplementary Figure 4 for "Plasma proteomics improves prediction of recurrent cardiovascular events"

**Supplementary Figure 4. Incremental predictive value of the combined model relative to SMART2 and the protein score for recurrent MACE. Forest plot of the change in C-index (ΔC-index) across testing datasets, with 95% confidence intervals. Blue indicates ΔC-indices relative to SMART2 alone when adding the protein score to SMART2; black indicates ΔC-indices relative to the protein score alone when adding SMART2 to the protein score. Positive values indicate incremental improvement of the combined model relative to the reference model within each testing dataset.**

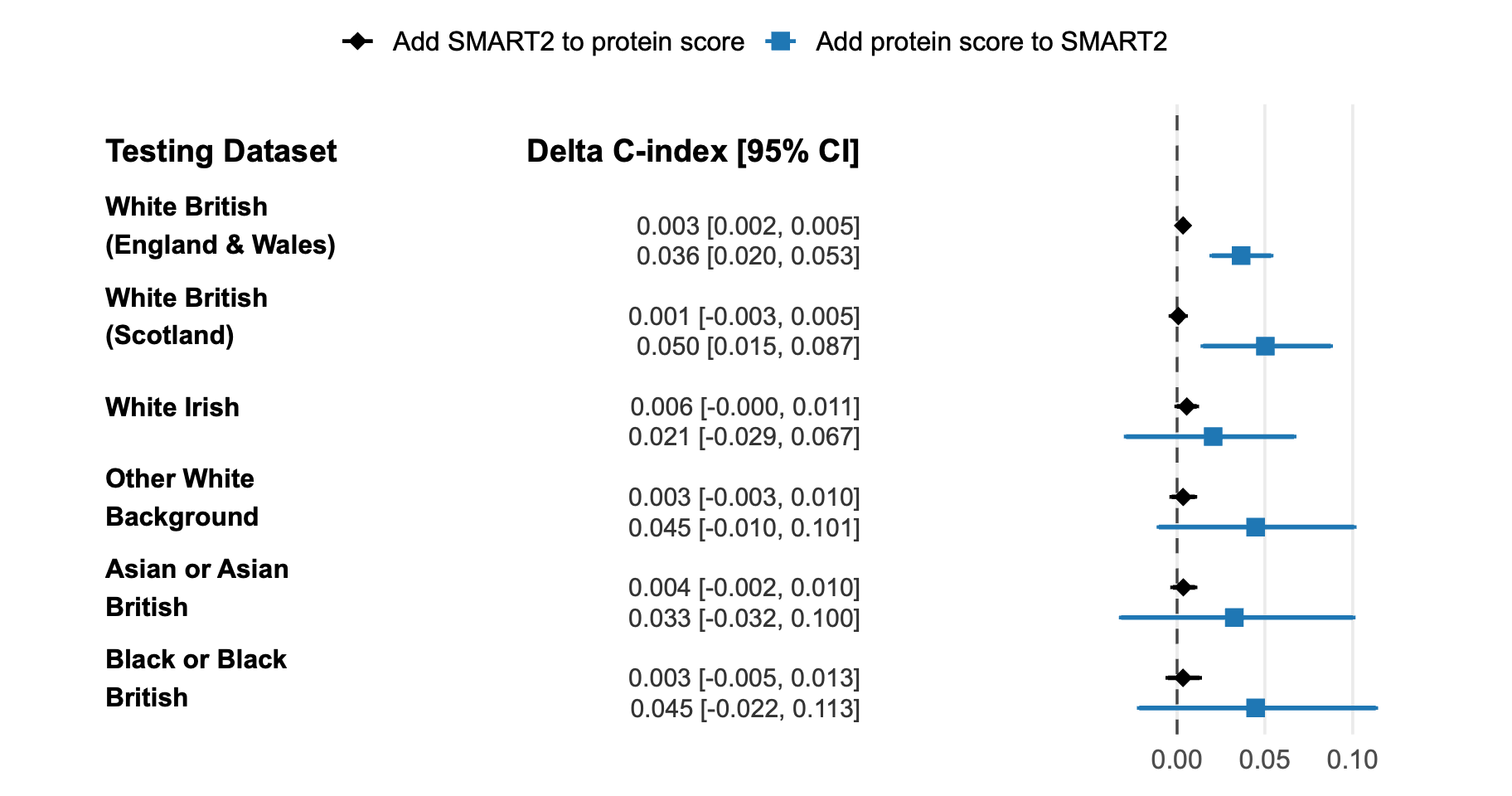
