## Supplementary Figure 5 for "Plasma proteomics improves prediction of recurrent cardiovascular events"

**Supplementary Figure 5. Incremental predictive value of the combined model relative to SMART2 and the 5-protein score. Forest plot of the change in C-index (ΔC-index) across testing datasets, with 95% confidence intervals. Blue indicates ΔC-indices relative to SMART2 alone when adding the 5-protein score to SMART2; black indicates ΔC-indices relative to the 5-protein score alone when adding SMART2. Positive values indicate incremental improvement of the combined model relative to the reference model within each testing dataset.**

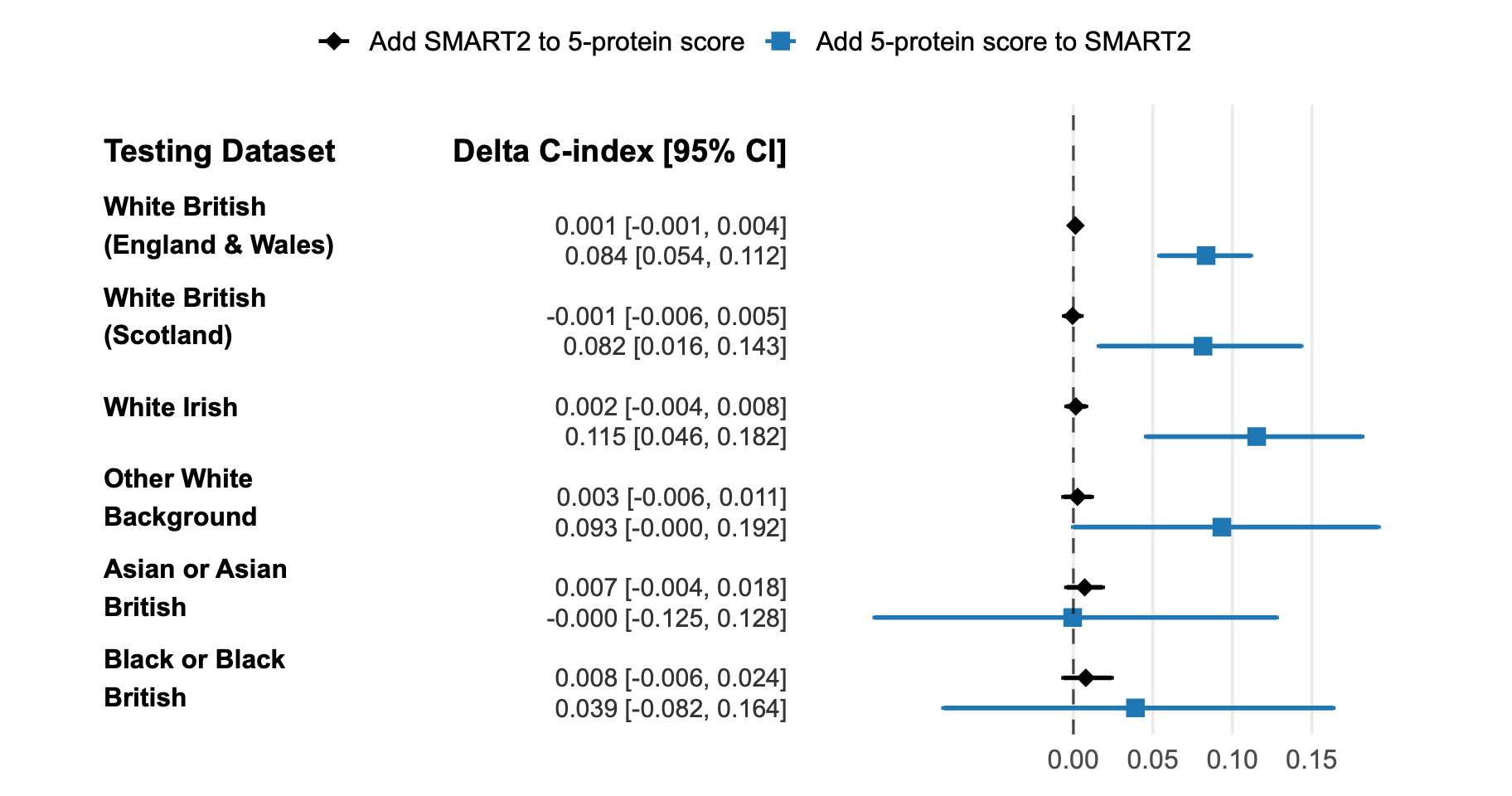
