## Supplementary Figure 6 for "Plasma proteomics improves prediction of recurrent cardiovascular events"

**Supplementary Figure 6. Comparative performance of the full protein score, the 5-protein score, and combined models for recurrent MACE across ethnic and geographic subgroups. Forest plot of Harrell’s C-index with 95% confidence intervals for the full protein score, the 5-protein score comprising growth differentiation factor 15 (GDF15), N-terminal pro-B-type natriuretic peptide (NT-proBNP), chromogranin A (CHGA), V-set and immunoglobulin domain-containing protein 2 (VSIG2), and angiotensin-converting enzyme 2 (ACE2), the SMART2 score, the age- and sex-adjusted 5-protein score, and the combined model integrating SMART2 with the 5-protein score across testing datasets.**

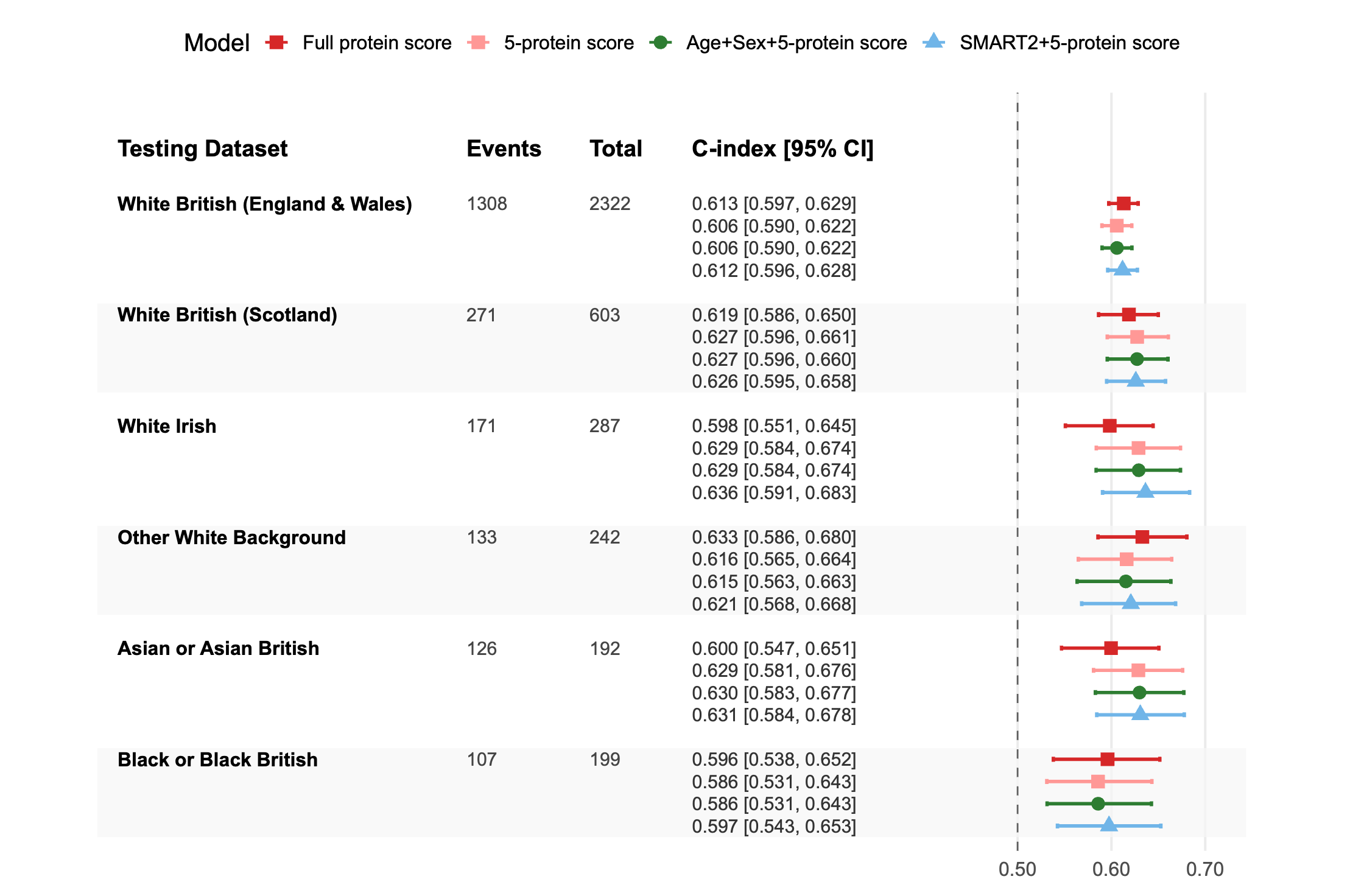
