## Supplementary Figure 7 for "Plasma proteomics improves prediction of recurrent cardiovascular events"

**Supplementary Figure 7. Incremental predictive value of the combined model relative to SMART2 and the 5-protein score for recurrent MACE. Forest plot of the change in C-index (ΔC-index) of the combined model relative to SMART2 alone (blue) or relative to the 5-protein score alone (black) across testing datasets. Positive values indicate better performance of the combined model relative to the reference model within that testing dataset.**

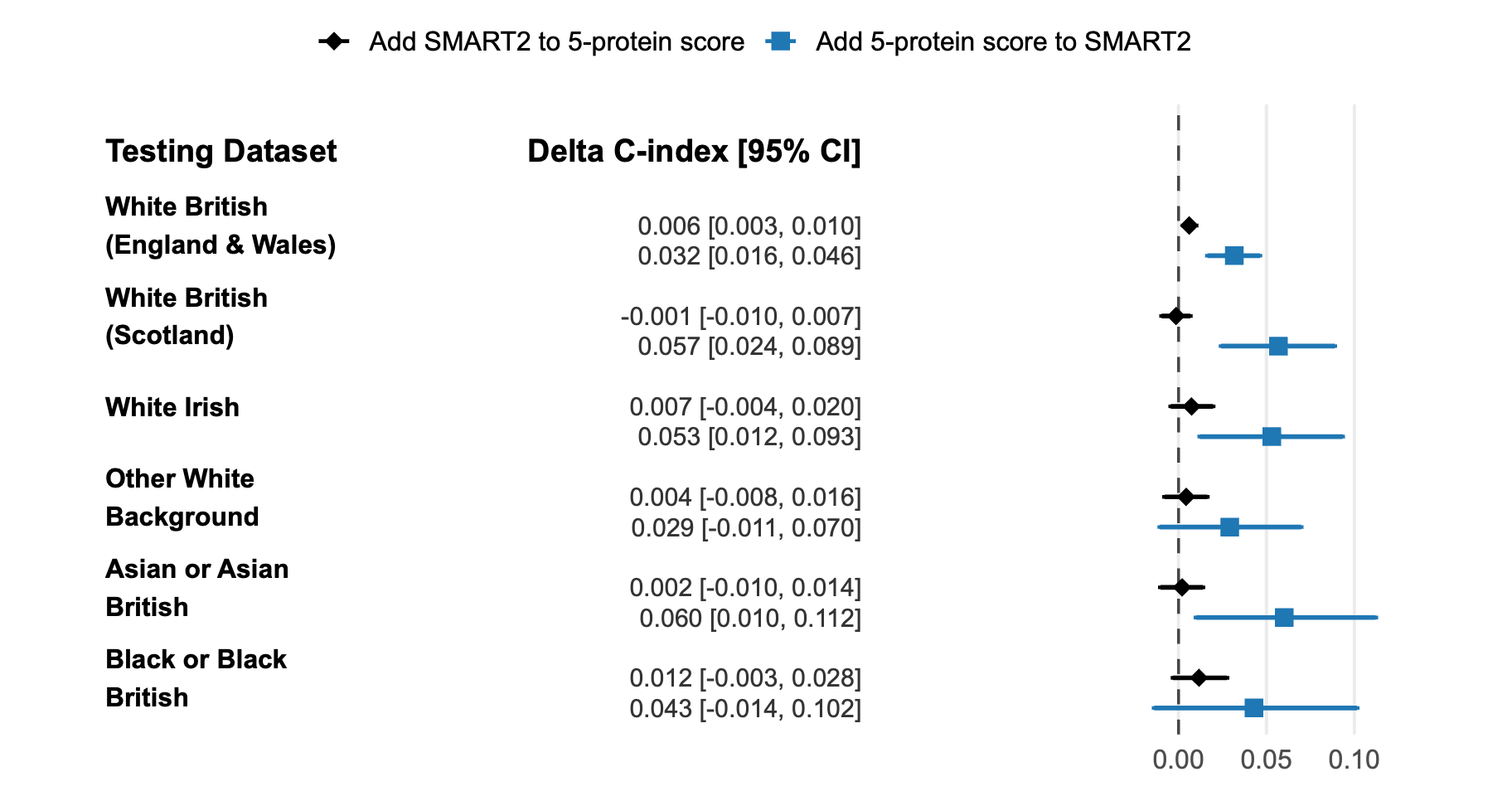
