## Supplementary Figure 8 for "Plasma proteomics improves prediction of recurrent cardiovascular events"

**Supplementary Figure 8. Calibration of predicted 10-year risk for recurrent ASCVD across testing datasets.** Calibration plots at a fixed 10-year horizon for the selected models in each testing dataset. Participants were grouped into deciles of predicted 10-year risk within each dataset. Points represent the mean predicted risk within each decile, plotted against the observed 10-year risk estimated using Kaplan–Meier methods; error bars indicate 95% confidence intervals for the observed risk.

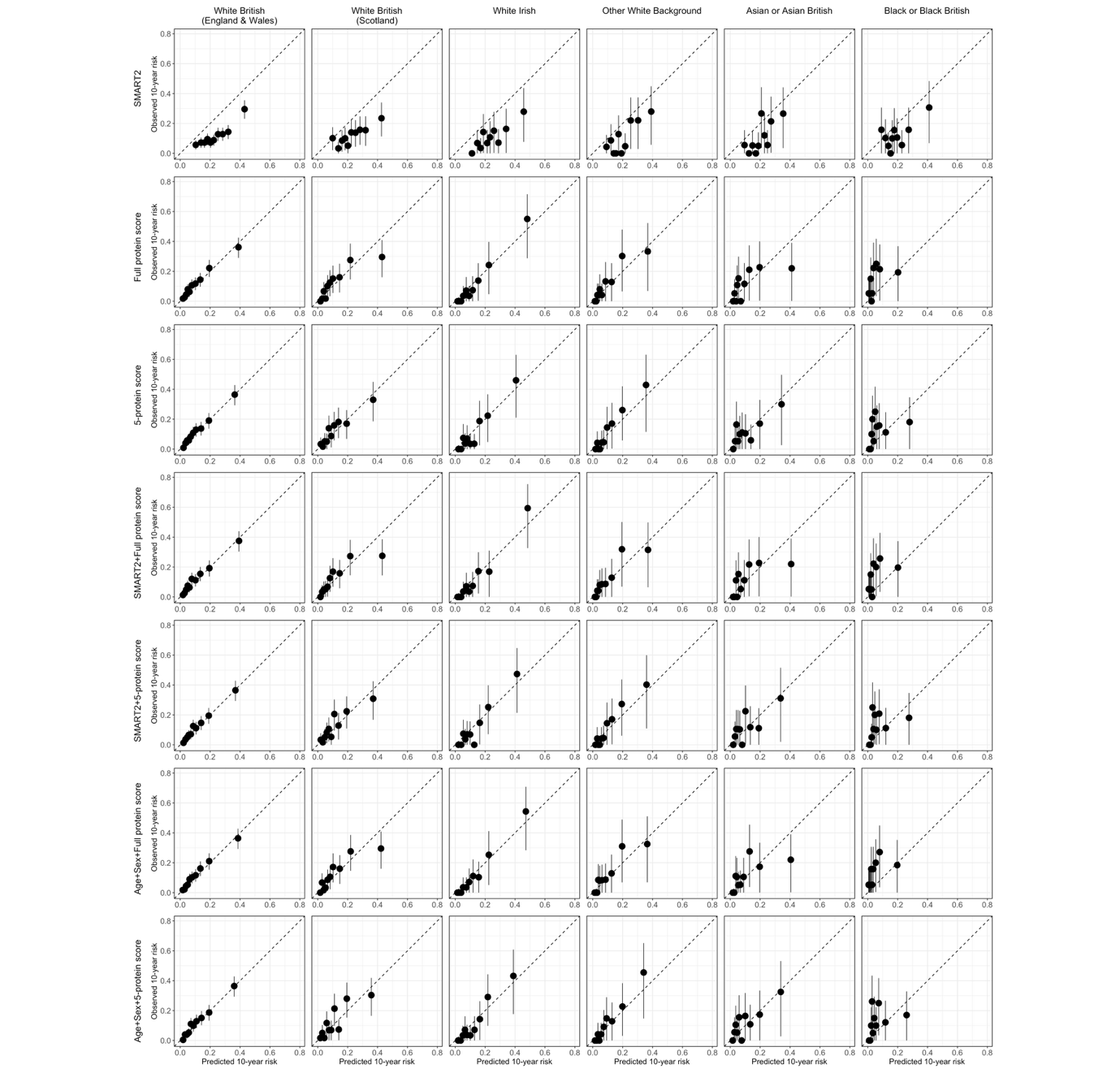
