## Supplementary Table 2 for "Plasma proteomics improves prediction of recurrent cardiovascular events"

**Supplementary Table 2.** Characteristics of UK Biobank Pharma Proteomics Project participants included in the present analysis of recurrent MACE (n = 9,356). Values are presented as median (interquartile range) for continuous variables and n (%) for categorical variables.

|  | Discovery cohort for training | Validation cohorts for testing | | | | | |
| --- | --- | --- | --- | --- | --- | --- | --- |
| Dataset | White British (England & Wales) | White British (England & Wales) | White British (Scotland) | White Irish | Other White Background | Asian or Asian British | Black or Black British |
| N | 5467 | 2342 | 605 | 294 | 251 | 197 | 200 |
| Length of follow-up, years | 13.49 (12.66–14.26) | 13.48 (12.64–14.27) | 14.82 (14.27–15.08) | 13.41 (12.56–14.35) | 13.42 (12.70–14.16) | 13.15 (12.57–13.90) | 13.12 (12.63–13.81) |
| Recurrent MACE, n (%) | 3083 (56.4%) | 1321 (56.4%) | 271 (44.8%) | 173 (58.8%) | 137 (54.6%) | 129 (65.5%) | 108 (54.0%) |
| Baseline age, years | 62.45 (56.75–66.44) | 62.37 (56.75–66.21) | 61.51 (55.27–66.43) | 62.37 (56.73–66.69) | 60.04 (53.32–65.22) | 57.61 (51.06–64.66) | 55.17 (49.53–62.78) |
| Female sex, n (%) | 2439 (44.6%) | 1014 (43.3%) | 283 (46.8%) | 140 (47.6%) | 130 (51.8%) | 86 (43.7%) | 118 (59.0%) |
| BMI, kg/m^2^ | 27.63 (24.91–30.91) | 27.52 (24.89–30.64) | 27.42 (25.00–30.54) | 27.33 (24.48–30.33) | 27.68 (24.15–30.91) | 27.37 (24.67–30.29) | 28.66 (25.83–31.89) |
| Systolic blood pressure (mmHg) | 138.0 (126.0–150.0) | 137.0 (126.0–150.0) | 139.0 (128.0–152.0) | 134.0 (124.0–146.8) | 133.0 (122.0–146.0) | 134.0 (122.0–146.0) | 137.0 (126.0–148.0) |
| Diastolic blood pressure (mmHg) | 81.0 (74.0–88.0) | 81.0 (74.0–88.0) | 82.0 (75.0–89.0) | 80.0 (72.0–86.0) | 79.0 (73.2–87.0) | 81.0 (74.0–88.0) | 83.0 (76.8–91.0) |
| Total cholesterol (mmol/L) | 5.22 (4.40–6.17) | 5.28 (4.46–6.14) | 5.15 (4.37–6.19) | 5.39 (4.41–6.23) | 5.24 (4.42–6.19) | 4.92 (4.03–5.60) | 5.02 (4.17–5.82) |
| HDL cholesterol (mmol/L) | 1.31 (1.09–1.56) | 1.31 (1.09–1.58) | 1.29 (1.06–1.58) | 1.36 (1.12–1.66) | 1.37 (1.16–1.61) | 1.16 (1.00–1.46) | 1.35 (1.15–1.64) |
| LDL cholesterol (mmol/L) | 3.20 (2.61–3.93) | 3.24 (2.65–3.92) | 3.18 (2.57–4.01) | 3.28 (2.51–3.85) | 3.22 (2.63–3.92) | 3.03 (2.36–3.66) | 3.04 (2.46–3.62) |
| Triglycerides (mmol/L) | 1.60 (1.14–2.29) | 1.62 (1.15–2.29) | 1.63 (1.13–2.42) | 1.51 (1.08–2.19) | 1.47 (1.09–2.17) | 1.66 (1.17–2.32) | 1.08 (0.80–1.47) |
| C-reactive protein (mg/L) | 1.65 (0.79–3.48) | 1.71 (0.80–3.45) | 1.76 (0.79–3.83) | 1.92 (0.85–4.20) | 1.38 (0.66–2.85) | 1.89 (0.84–3.39) | 1.69 (0.71–3.23) |
| Creatinine (μmol/L) | 73.8 (63.5–85.3) | 74.4 (64.0–86.4) | 72.1 (61.3–83.2) | 73.5 (62.3–83.3) | 71.8 (62.7–84.1) | 69.8 (60.3–80.5) | 77.1 (67.1–89.3) |
| Current smoking, n (%) | 576 (10.5%) | 269 (11.5%) | 93 (15.4%) | 63 (21.4%) | 32 (12.7%) | 18 (9.1%) | 23 (11.5%) |
| Diabetes mellitus, n (%) | 559 (10.2%) | 228 (9.7%) | 58 (9.6%) | 24 (8.2%) | 38 (15.1%) | 60 (30.5%) | 35 (17.5%) |
| Antithrombotic medication, n (%) | 2169 (39.7%) | 909 (38.8%) | 276 (45.6%) | 114 (38.8%) | 99 (39.4%) | 93 (47.2%) | 51 (25.5%) |
